# Spatial Patterns of COVID-19 Mortality: Examining Socioeconomic Determinants in U.S. Counties Using Cluster Analysis

**DOI:** 10.1101/2024.10.30.24316443

**Authors:** Robert Ross Wardrup

## Abstract

**Aim:** This study aims to investigate the spatial patterns of COVID-19 mortality across U.S. counties and identify the socioeconomic determinants that influence these mortality trends, using spatial epidemiological methods.

**Subject and Methods:** We conducted a spatial analysis of COVID-19 mortality data from over 3,000 U.S. counties, applying cluster detection techniques, including SatScan, to identify areas with significant mortality trends. Spatial regression models, including spatial lag and spatial error models, were employed to examine the impact of socioeconomic variables, such as race, income inequality, and insurance rates, on COVID-19 mortality. The analysis controlled for multicollinearity and spatial autocorrelation in the data.

**Results:** Counties with higher proportions of Black populations and higher uninsured rates exhibited significantly lower COVID-19 trends over the study period. Spatial clustering revealed regions in the northwestern and eastern/northeastern United States with a mix of positive and negative mortality rate trends. The spatial lag model showed the strongest fit, confirming the importance of spatial dependency in explaining mortality patterns.

**Conclusion:** This study highlights the significant spatial disparities in COVID-19 mortality across U.S. counties. The findings emphasize the need for targeted public health interventions in vulnerable regions to address these disparities.

## Introduction

The COVID-19 pandemic has had catastrophic global effects, influencing public health, economic stability, and social structures. In the United States, the unemployment rate surged from 10.3% to 14.7% between March and April 2020. The U.S. healthcare system faced immense strain, with only 2.9 hospital beds per 1,000 people and 34.7 ICU beds per 100,000 people (Shrestha et al., 2020). Moreover, the pandemic disproportionately impacted different populations, exacerbating pre-existing social, racial, and economic disparities (Yancy, 2020).

Numerous studies have highlighted how these disparities contributed to worsened COVID-19 outcomes. Hawkins et al. (2020) found that counties with lower educational attainment, higher Black populations, and greater poverty rates experienced higher COVID-19 mortality. Similarly, Millett et al. (2020) demonstrated that Black Americans more likely to die from COVID-19 compared to White Americans, due in part to structural inequalities, including access to healthcare and economic disparities. Polyakova et al. (2021) further reinforced the role of socioeconomic inequalities by showing that states like Michigan and Louisiana, which have larger Black populations, experienced disproportionately high mortality rates.

Although these studies provide valuable insights, relatively few have employed spatial methods to investigate the relationship between socioeconomic status (SES) and COVID-19 mortality, especially at the county level. Spatial patterns of disease provide essential context for understanding how regional factors influence health outcomes. Studies using spatial epidemiological methods, such as Desjardins, Hohl, and Delmelle (2020), have shown geographic clusters of high COVID-19 incidence. Additionally, Kang et al. (2020) employed spatial-temporal analysis in China to identify regions with rapidly increasing COVID-19 cases. Despite these important findings, few studies have applied spatial cluster analysis specifically to COVID-19 mortality, particularly in the United States.

To address this gap, we use SatScan’s Spatial Variations in Temporal Trends (SVTT) analysis to identify clusters of U.S. counties with statistically significant COVID-19 mortality trends. SatScan is a statistical software designed for the detection of disease outbreaks by scanning for clusters of disease cases or deaths in space and time. The method compares the observed cases within a defined geographic area to the expected number of cases, based on population size and other variables, identifying areas where mortality or case counts significantly deviate from what is expected (Kulldorff et al., 1997). The SVTT module in SatScan specifically identifies clusters where mortality trends are increasing or decreasing over time, which allows for the detection of regions with particularly concerning or improving trends. By applying SaTScan, we isolate regions of interest while avoiding noise from random fluctuations, ensuring that our analysis focuses on counties exhibiting meaningful spatial-temporal patterns.

Previous research has demonstrated that spatial analysis techniques such as SaTScan can be valuable for identifying and understanding the spread of disease. For example, Desjardins et al. (2020) used SatScan to track COVID-19 transmission dynamics in the U.S. Other studies, such as Gross et al. (2021), have also used spatial methods to map the unequal burden of COVID-19 on minority populations, reinforcing the importance of spatial techniques for public health interventions.

This study aims to explore significant clusters of COVID-19 mortality trends and the socioeconomic attributes driving these disparities, offering a focused analysis of regions with statistically significant mortality trends. Spatial regression models, including spatial lag and spatial error models, are then employed to account for spatial dependence in the data, further refining our understanding of the role that socioeconomic conditions and regional factors play in shaping mortality outcomes during public health crises.

## Methodology

### Data Collection & Sources

The data for this analysis were sourced from the U.S. Census Bureau’s American Community Survey (ACS) and the Centers for Disease Control and Prevention (CDC). The socioeconomic and demographic data were obtained using ACS 5-year estimates for 2020 at the county level, while COVID-19 mortality data were gathered from the CDC’s National Vital Statistics System for the years 2019 to 2022. The goal was to explore the relationship between socioeconomic status (SES) indicators and COVID-19 mortality trends in U.S. counties.

### COVID-19 Mortality Data Processing

The COVID-19 mortality data were aggregated at the county level. The data were cleaned by replacing suppressed data values with missing values (NA) and converting the reported deaths into integers. Counties with missing or suppressed death counts were excluded from the analysis to avoid introducing bias or uncertainty. While imputation techniques could have been considered, exclusion was chosen to maintain the integrity of the spatial patterns and avoid potential distortions in mortality trends due to unreliable data.

The analysis was limited to counties identified as part of spatial clusters using SatScan’s SVTT method, which isolates regions with statistically significant increases or decreases in mortality over time. This approach enhances the precision of the study by excluding counties without significant patterns, thereby reducing noise and improving the detection of meaningful associations between socioeconomic factors and mortality trends.

SatScan’s Spatial Variations in Temporal Trends (SVTT) test was employed to detect clusters where COVID-19 mortality trends were either increasing or decreasing over time. By restricting the analysis to these clusters, we ensured that the study was concentrated on counties that showed statistically significant patterns in mortality, thereby improving the ability to detect meaningful relationships between socioeconomic variables and mortality trends. This approach allowed us to avoid including counties with random or non-significant trends, which could dilute the findings and obscure important relationships.

The default Population At Risk (PAR) setting in SaTScan allows up to 20% of the population at risk to be included in each cluster. However, when applying this default to the current study, the clusters identified were too large, often encompassing multiple regions of the continental United States. Large clusters can be problematic because they reduce the granularity of the analysis, making it difficult to detect localized variations in COVID-19 mortality trends and their associations with socioeconomic factors. Large, multi-region clusters might obscure the more precise, smaller-scale relationships that exist within specific counties or smaller regions.

To address this, an iterative process was undertaken, gradually reducing the PAR parameter until cluster sizes were more appropriate for the study’s objectives. Smaller clusters offer a higher level of specificity, allowing for a more accurate exploration of the associations between SES variables and COVID-19 mortality trends within distinct geographic areas. This adjustment ensures that the study captures local-level trends and avoids the loss of statistical power that could result from overly broad spatial clusters.

In the study, a Standard Monte Carlo approach with 999 replications was employed to calculate p-values for the significance of detected clusters. Monte Carlo simulations are a robust method to assess statistical significance in spatial analysis, particularly in SaTScan, where cluster detection relies on random simulations to establish the distribution of the test statistic under the null hypothesis (Kulldorff, et al., 1997).

The choice of 999 replications is standard practice in spatial epidemiology, as it strikes a balance between computational efficiency and the accuracy of p-value estimation. This number of replications ensures that the simulation provides a stable and reliable distribution of the log-likelihood ratio (LLR) test statistic, from which the statistical significance of the clusters can be determined. The use of Monte Carlo simulations helps minimize the risk of type I errors (false positives) when detecting clusters, ensuring that the findings are statistically robust and not due to random chance.

By adjusting for more likely clusters and applying a cut-off p-value of 0.05, we ensured that only clusters with a high degree of confidence in their statistical significance were included in the analysis. This method provides additional robustness to the findings, ensuring that the reported clusters reflect true spatial patterns in COVID-19 mortality trends.

We aggregated the data by three-month intervals due to issues with model convergence when using shorter time periods. Monthly data, while more granular, introduced challenges related to model stability and computation. By aggregating to a quarterly basis, we were able to smooth out short-term fluctuations and ensure more reliable convergence in the spatial regression models, ultimately improving the robustness of the analysis. This approach also captures longer-term trends in COVID-19 mortality without losing the temporal dimension necessary for the analysis. The final SaTScan parameters are shown in table 2.

**Table 1:** SatSCan Parameters Used.

| Parameter | Setting |
| --- | --- |
| Type of Analysis | Spatial Variation in Temporal Trends |
| Probability Model | Discrete Poisson |
| Scan for Areas with | Increasing or Decreasing Rates |
| Time Aggregation Units | Month |
| Time Aggregation Length | 3 |
| Maximum Spatial Cluster Size | 0.5 percent of population at risk |
| Window Shape | Circular |
| Minimum Cases in Cluster for High Rates | 2 |
| Restrict High Rate Clusters | No |
| Restrict Low Rate Clusters | No |
| Temporal Adjustment | None |
| P-Value Reporting | Standard Monte Carlo |
| **Number of Replications** | 999 |
| **Adjusting for More Likely Clusters** | Yes |
| **Maximum number of iterations** | 10 |
| **Stop when p-value greater** | 0.05 |
| **Standard Drilldown on Detected Clusters** | Yes |
| **Minimum Locations in Detected Cluster** | 2 |
| **Minimum Cases in Detected Cluster** | 10 |
| **Cutoff of Detected Cluster** | 0.05 |
| **Report Hierarchical Clusters** | Yes |
| **Criteria for Reporting Secondary Clusters** | No Geographical Overlap |
| **Restrict Reporting to Smaller Clusters** | No |
| **Report Critical Values** | Yes |
| **Report Monte Carlo Rank** | Yes |

**Table 2:** Socioeconomic Variable Descriptive Statistics.

| **Variable** | **Mean** | **Median** | **Min** | **Max** | **Std. Dev.** |
| --- | --- | --- | --- | --- | --- |
| **Median Age** | 43.63 | 43.7 | 30 | 56.3 | 3.953 |
| **Median Income** | \$49,189.00 | \$48,350.00 | \$22,292.00 | \$107,246.00 | \$12,657.50 |
| **Poverty Rate** | 16.97% | 16.13% | 4.59% | 36.10% | 6.14% |
| **Unemployment Rate** | 6.30% | 5.72% | 1.70% | 25.43% | 2.65% |
| **Uninsured Rate** | 0.15% | 0.10% | 0.00% | 1.15% | 0.18% |
| **Overcrowding Rate** | 0.79% | 0.06% | 0.00% | 0.86% | 0.89% |
| **Public Transport Usage Rate** | 0.34% | 0.10% | 0.00% | 10.55% | 1.13% |
| **Percent Black** | 4.97% | 1.70% | 0.00% | 50.07% | 8.77% |
| **Percent Asian** | 0.96% | 0.54% | 0.00% | 16.53% | 1.51% |
| **Percent Hispanic** | 3.83% | 1.83% | 0.00% | 31.95% | 4.70% |

**Table 3:** Significant SaTScan SVTT Clusters & RR.

| Cluster | Counties | P-Value | Obs. Mortality | Exp. Mortality | RR |
| --- | --- | --- | --- | --- | --- |
| 1 | 66 | 0.001 | 7185 | 3903.91 | 1.85 |
| 2 | 3 | 0.001 | 4693 | 3766.95 | 1.25 |
| 3 | 2 | 0.001 | 5216 | 3729.88 | 1.4 |
| 4 | 28 | 0.001 | 3602 | 3786.85 | 0.95 |
| 5 | 59 | 0.001 | 5467 | 3802.46 | 1.44 |
| 6 | 1 | 0.001 | 3019 | 2977.41 | 1.01 |
| 7 | 25 | 0.001 | 2550 | 3120.83 | 0.82 |
| 8 | 35 | 0.001 | 6365 | 3164.84 | 2.02 |
| 9 | 18 | 0.001 | 5970 | 3135.35 | 1.91 |
| 10 | 21 | 0.001 | 5162 | 3585.74 | 1.44 |

**Table 4:**
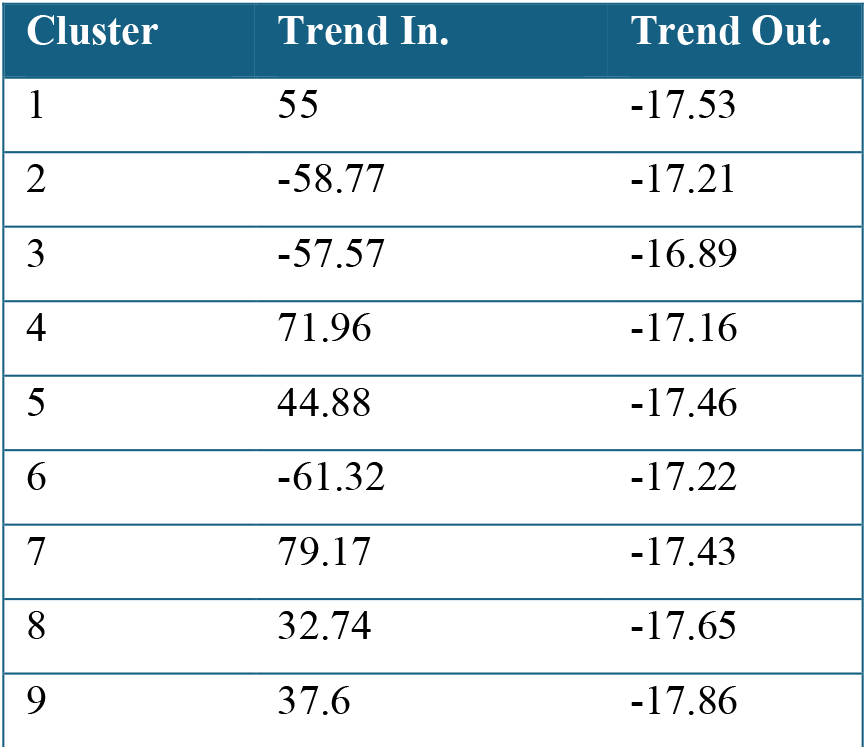
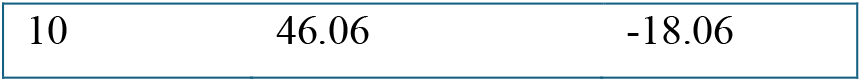
Significant SaTScan SVTT Clusters & Mortality Trends.

**Table 5:** Ordinary Least Squares Model Results.

| Variable | Estimate | Std. Error | t value | Pr(>\|t\|) |
| --- | --- | --- | --- | --- |
| (Intercept) | 128 | 130.9 | 0.978 | 0.32906 |
| median_age | -1.148 | 1.817 | -0.632 | 0.52812 |
| median_income | -4.71E-06 | 0.001028 | -0.005 | 0.99635 |
| poverty_rate | -1.592 | 2.145 | -0.742 | 0.4587 |
| unemployment_rate | 5.989 | 3.618 | 1.656 | 0.09937 |
| uninsured_rate | -91.72 | 33.76 | -2.717 | 0.00716 |
| overcrowding_rate | 145.2 | 79.41 | 1.828 | 0.06901 |
| public_transport_usage_rate | -17.73 | 7.532 | -2.354 | 0.01953 |
| race_black_rate | -1.817 | 0.7166 | -2.536 | 0.01199 |
| race_asian_rate | 7.706 | 5.947 | 1.296 | 0.19652 |
| hispanic_rate | -2.107 | 1.757 | -1.199 | 0.23191 |
| AIC | 2470.1 | N/A | N/A | N/A |
| Moran's I of Residuals | 0.125 (Exp.: -0.005) | N/A | Z=2.762 | 0.003 |

### SES Data & Variables

To capture socioeconomic and demographic profiles of each county, we extracted variables from the 2020 ACS 5-year estimates, which included:

- **Median income**
- **Poverty rate**
- **Unemployment rate**
- **Percent with no health insurance**
- **Overcrowding rate**: The percentage of households with more than 1.5 occupants per room.
- **Public transportation usage**: The percentage of people using public transportation.
- **Race/ethnicity variables**: The percentage of the population identifying as White, Black or African American, Asian, or Hispanic or Latino.
- **Median age**

Each SES variable was calculated as a percentage of the total population in each county where applicable. For example, the unemployment rate was calculated by dividing the number of unemployed individuals by the total labor force in each county.

### Identifying Spatial Clusters

Spatial clusters of counties with similar COVID-19 mortality trends were identified using SaTScan’s Spatial Variations in Temporal Trends analysis. This method identifies spatial and temporal clusters of events, such as disease cases or deaths, by detecting areas where the observed number of cases significantly exceeds what is expected. It is used to find clusters of unusually high or low event rates in both space and time. Counties that were detected as part of a cluster were included in this analysis, while counties not identified as part of a cluster were excluded from trend-based modeling.

The decision to exclude counties not identified as part of spatial clusters in this research is grounded in the methodology of SaTScan’s SVTT model, which detects areas with statistically significant clusters of mortality trends. Counties outside of these clusters were not flagged as having unusual patterns in the data and including them could dilute the statistical power and focus of the analysis. By concentrating only on counties with significant spatial or temporal mortality trends, this research aims to better understand the factors influencing these distinct regions and avoid introducing noise from areas with no significant trends.

### Data Preparation & Aggregation

SES metrics were calculated using American Community Survey (ACS) data and merged with COVID-19 mortality data from the CDC by matching county geographic identifiers (GEOIDs). To minimize the impact of short-term fluctuations, mortality data were aggregated at three-month intervals, smoothing out seasonal or monthly variations and ensuring model convergence in subsequent spatial regression analyses.

### Statistical Models

We used several regression models to examine the relationship between SES indicators and the COVID-19 mortality trends in counties within identified spatial clusters.

1. **Ordinary Least Squares (OLS) Model**: Initially, a linear regression model was fitted using COVID-19 mortality trends as the dependent variable and the SES indicators as independent variables. Multicollinearity was assessed using variance inflation factors (VIF), with a threshold of VIF > 10 indicating problematic multicollinearity. High collinearity was observed between race_white_rate and race_black_rate. After assessing their respective importance to the model, race_white_rate was excluded to retain race_black_rate, as previous literature suggests that the Black population has been disproportionately affected by COVID-19 mortality. This decision was based on both statistical considerations and substantive relevance to the study.
2. **Spatial Lag Model**: To account for spatial dependence in the data, we employed a spatial lag model. This model assumes that the mortality trend in each county is influenced by the trends in neighboring counties, with spatial weights determined by contiguity. The inclusion of a spatial lag variable helps correct for spatial autocorrelation in the data.
3. **Spatial Error Model**: A spatial error model was used as an alternative to the spatial lag model. This model corrects for spatial autocorrelation by accounting for spatial dependence in the error terms of the regression model.
4. **Models with Interaction Terms**: To explore potential interactions between SES variables, we included interaction terms in the spatial models. Specifically, we examined interactions between median income and race_black_rate, as well as other combinations of SES indicators such as unemployment rate and insurance coverage.

### Spatial Weights & Autocorrelation

A queen contiguity spatial weights matrix, which defines neighboring counties based on shared borders, was applied in both the spatial lag and spatial error models to capture spatial dependence. The Moran’s I statistic was computed to assess spatial autocorrelation in the OLS model’s residuals, and the significant results (Moran’s I = 0.125, p=0.003) justified the need for spatial econometric models.

#### Model Selection

The spatial lag model without interaction terms was selected as the best-performing model based on the lowest Akaike Information Criterion (AIC) value and its ability to mitigate residual spatial autocorrelation, providing a better fit for the data than the OLS or spatial error models.

## Results

### SaTScan Results

The SaTScan SVTT analysis identified 10 statistically significant clusters of counties across the continental United States, highlighting regions with notable variations in COVID-19 mortality trends over time. Cluster 1, the largest cluster, comprised 66 counties with a log likelihood ratio (LLR) of 891.92 and a relative risk (RR) of 1.85, indicating that mortality in these counties was 85% higher than expected. This cluster recorded 7,185 observed deaths, significantly exceeding the 3,903.91 expected deaths. The “Trend In” value of 55 indicated that, despite the high mortality, the trend within this cluster had been increasing over the study period, suggesting a worsening mortality rate. In contrast, the “Trend Out” value of -17.53 showed that mortality rates were declining outside this cluster, emphasizing the need for targeted interventions in these counties.

Similarly, Cluster 8, with 35 counties, had the highest relative risk (RR=2.02), suggesting that mortality rates in these counties were more than twice as high as expected. The cluster recorded 6,365 observed deaths compared to 3,164.84 expected deaths. The “Trend In” value of 32.74 suggested an increasing mortality trend within the cluster, while the “Trend Out” value of -17.65 reflected a decrease in mortality trends outside of it. This contrast pointed to regions that had ongoing mortality challenges, despite improvements elsewhere.

In contrast, some clusters, such as Cluster 4 and Cluster 7, exhibited relative risks below 1, indicating lower-than-expected mortality rates. For example, Cluster 7, composed of 25 counties, had a relative risk of 0.82, with 2,550 observed deaths compared to the expected 3,120.83. The “Trend In” value of 79.17 indicated a sharp increase in mortality over the study period within the cluster, while the “Trend Out” value of -17.43 suggested a decrease outside the cluster. Despite lower-than-expected deaths overall, this cluster warranted attention due to its rising mortality trend.

The results from this analysis emphasized the geographic disparities in COVID-19 mortality trends, with some clusters experiencing rising mortality rates over the study period, while others saw declines. This highlighted the need for targeted interventions in regions that showed increasing mortality rates, while acknowledging the success of efforts in areas with decreasing trends. Including the “Trend In” and “Trend Out” values provided a more nuanced understanding of how mortality rates evolved both within and outside each cluster.

**Figure 1.**
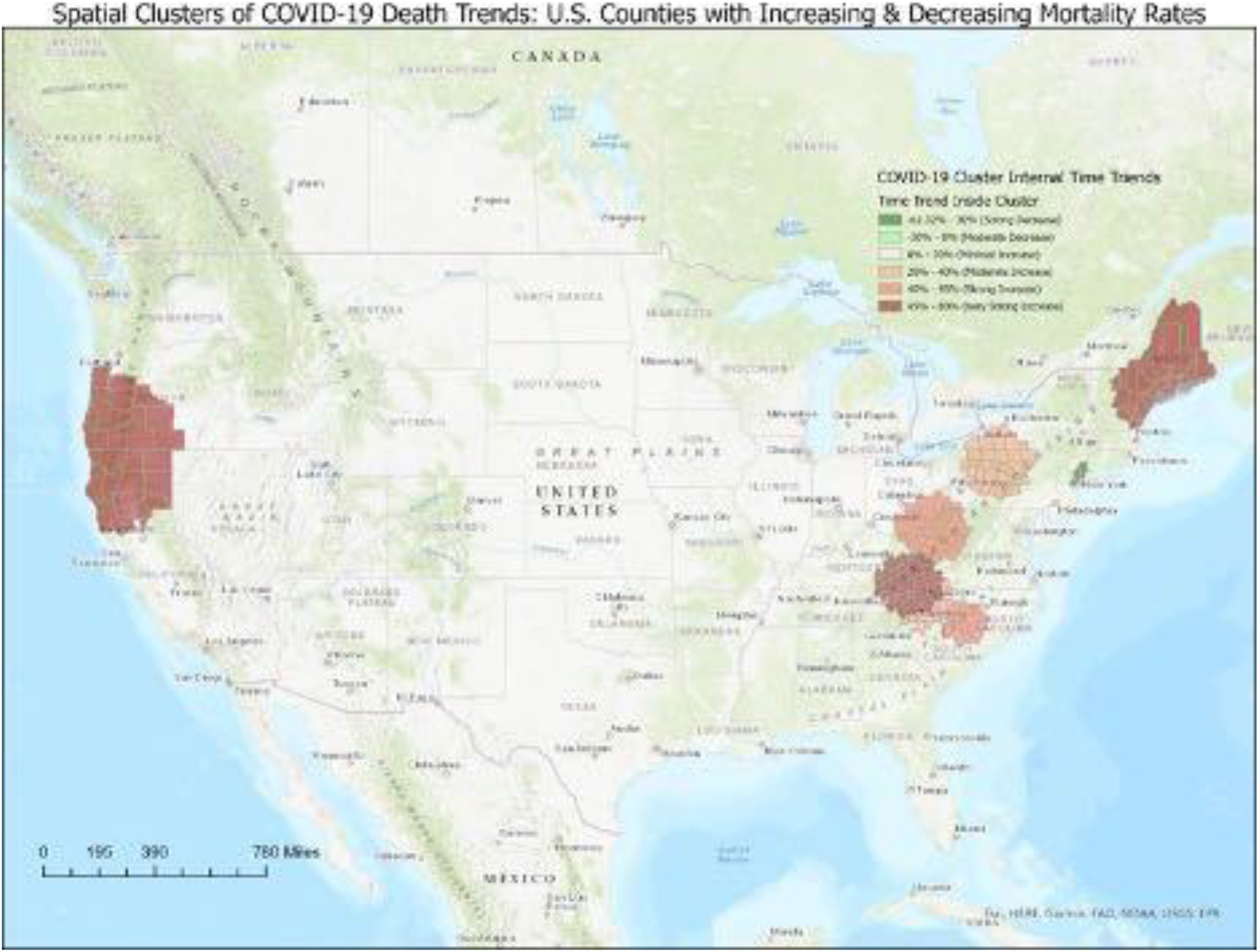
Internal Mortality Rate Trends of Significant SaTScan SVTT Clusters.

### OLS Regression Results (Main Effects Model)

We first estimated an ordinary least squares (OLS) regression model with mortality trend (LOC_TRENDLOC) as the dependent variable and key SES indicators as independent variables. The SES indicators included median age, median income, poverty rate, unemployment rate, percent without medical insurance, overcrowding rate, public transport usage rate, and racial/ethnic population proportions (Black, Asian, and Hispanic rates).

The OLS model was statistically significant (p < 0.001, table 8) and revealed statistically significant associations between several SES variables and COVID-19 mortality trends. Counties with higher uninsured rates (β=−91.72, p=0.00716) and higher public transport usage rates (β=−17.73, p=0.01953) showed more negative mortality trends, indicating a decrease in mortality over time. A higher proportion of Black residents was also significantly associated with more negative trends (β=−1.82,p=0.01199). These findings were counterintuitive, particularly for the uninsured rate, which may reflect delayed healthcare access, late-stage public health interventions, or timing differences in pandemic waves. The adjusted R^2^ of the model was 0.1237, indicating that about 12% of the variation in mortality trends could be explained by these SES variables.

Despite these results, the model diagnostics raised concerns about spatial autocorrelation in the residuals. A Moran’s I test revealed significant spatial autocorrelation (Moran’s I = 0.125, p=0.003), suggesting that an OLS regression may not fully capture the spatial dependence present in the data.

### Spatial Lag Model

To address spatial dependence, which was confirmed by significant spatial autocorrelation in the OLS residuals, a spatial lag model was employed. This model assumes that the mortality trend in each county is not independent of neighboring counties, with spatial relationships modeled through a queen contiguity spatial weights matrix. The spatial lag parameter (ρ=0.27275, p=0.001) was statistically significant, further justifying the use of a spatial econometric approach. This model improved the fit compared to OLS by successfully addressing residual spatial autocorrelation, as indicated by the reduction in the Lagrange Multiplier statistic (LM=3.251, p=0.071).

The spatial lag model was statistically significant (p=0.001, table 8) and indicated that the uninsured rate (β=−82.80,p=0.0095), and Black population rate (β=−1.52,p=0.027) were negatively associated with mortality trends. However, the magnitude of the coefficients was somewhat reduced compared to the OLS model. The AIC of the spatial lag model was 2462.1, the lowest of the spatial models. The Lagrange Multiplier test for residual autocorrelation was not statistically significant (LM=3.251, p=0.071), indicating that the spatial lag model successfully addressed spatial autocorrelation.

### Spatial Error Model

An alternative spatial error model was also estimated to account for potential spatial error dependence. This model was statistically significant (p < 0.001, table 8). In the error model, the lambda parameter (λ=0.28315,p=0.005) was statistically significant, indicating that spatial error dependence was present. The findings from the spatial error model were largely consistent with those from the spatial lag model. Uninsured rate (β=−90.82,p=0.006) and Black population rate (β=−1.88,p=0.021) remained significant predictors of negative mortality trends.

### Interaction Models

Finally, interaction terms were added to the spatial models to test whether the effects of SES variables on COVID-19 mortality trends varied across different racial or economic contexts. Specifically, interactions between median income and race Black rate, as well as median income and poverty rate, were included in the models.

The spatial lag model was statistically significant (p < 0.001, table 8). Both race_black_rate and uninsured rate were found to be statistically significant predictors. The interaction between median income and race Black rate was not statistically significant (β=−0.42, p=0.951), nor were any of the other interaction terms (Table 6). Similarly, in the spatial error model with interaction terms, no interactions were found to be significant.

**Table 6:**
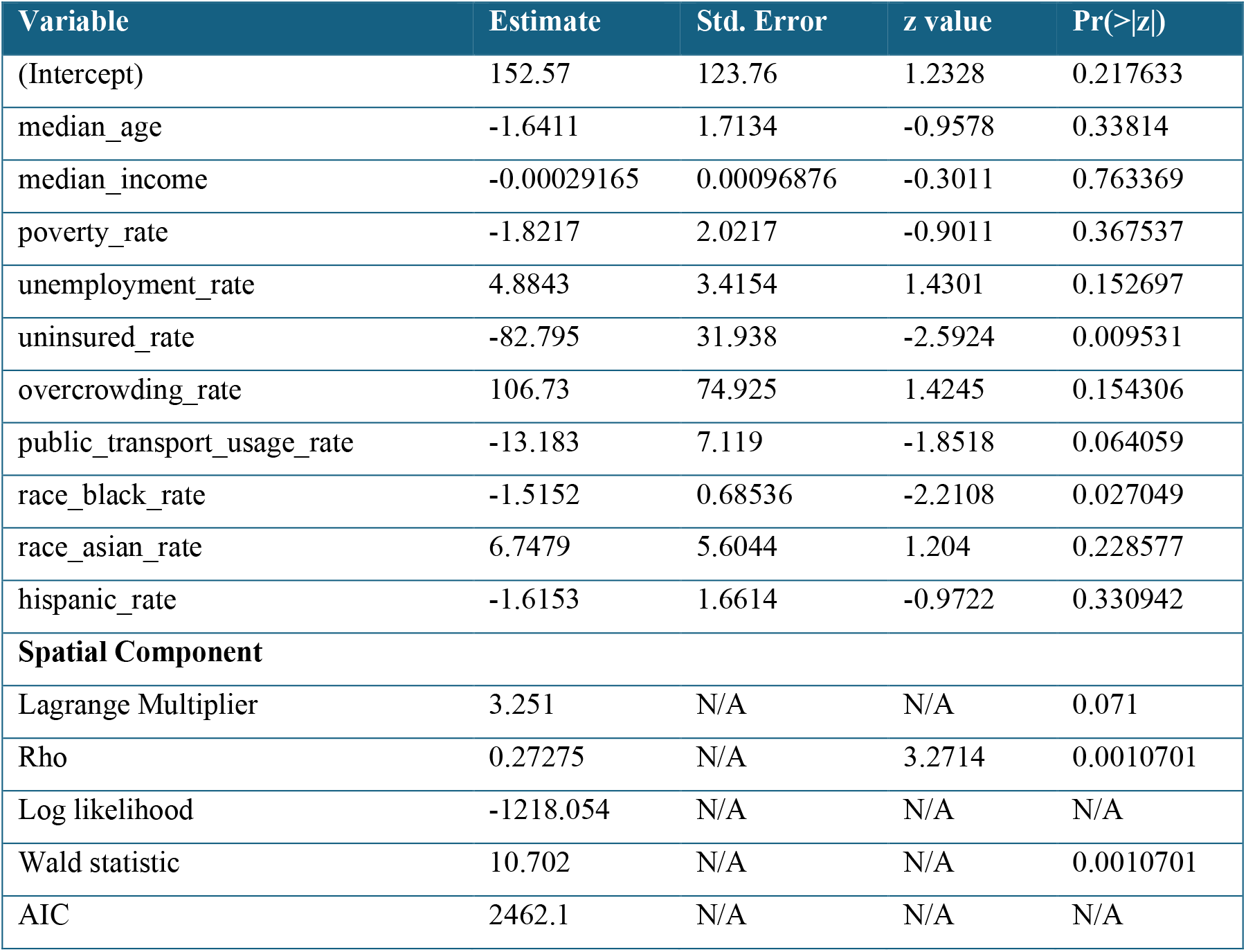
Spatial Lag Model Results.

| Variable | Estimate | Std. Error | z value | Pr(>\|z\|) |
| --- | --- | --- | --- | --- |
| (Intercept) | 152.57 | 123.76 | 1.2328 | 0.217633 |
| median_age | -1.6411 | 1.7134 | -0.9578 | 0.33814 |
| median_income | -0.00029165 | 0.00096876 | -0.3011 | 0.763369 |
| poverty_rate | -1.8217 | 2.0217 | -0.9011 | 0.367537 |
| unemployment_rate | 4.8843 | 3.4154 | 1.4301 | 0.152697 |
| uninsured_rate | -82.795 | 31.938 | -2.5924 | 0.009531 |
| overcrowding_rate | 106.73 | 74.925 | 1.4245 | 0.154306 |
| public_transport_usage_rate | -13.183 | 7.119 | -1.8518 | 0.064059 |
| race_black_rate | -1.5152 | 0.68536 | -2.2108 | 0.027049 |
| race_asian_rate | 6.7479 | 5.6044 | 1.204 | 0.228577 |
| hispanic_rate | -1.6153 | 1.6614 | -0.9722 | 0.330942 |
| **Spatial Component** |  |  |  |  |
| Lagrange Multiplier | 3.251 | N/A | N/A | 0.071 |
| Rho | 0.27275 | N/A | 3.2714 | 0.0010701 |
| Log likelihood | -1218.054 | N/A | N/A | N/A |
| Wald statistic | 10.702 | N/A | N/A | 0.0010701 |
| AIC | 2462.1 | N/A | N/A | N/A |

**Table 7:** Spatial Error Model Results.

| Variable | Estimate | Std. Error | z value | Pr(>\|z\|) |
| --- | --- | --- | --- | --- |
| (Intercept) | 249.33 | 129.17 | 1.9303 | 0.05357 |
| median_age | -2.4483 | 1.7958 | -1.3634 | 0.172762 |
| median_income | -0.00078985 | 0.0010569 | -0.7473 | 0.454859 |
| poverty_rate | -2.3317 | 2.1551 | -1.082 | 0.279272 |
| unemployment_rate | 4.4988 | 3.6216 | 1.2422 | 0.214158 |
| uninsured_rate | -90.817 | 33.098 | -2.7439 | 0.006072 |
| overcrowding_rate | 76.198 | 77.643 | 0.9814 | 0.326398 |
| public_transport_usage_rate | -11.802 | 7.6341 | -1.5459 | 0.122121 |
| race_black_rate | -1.8813 | 0.81646 | -2.3043 | 0.021208 |
| race_asian_rate | 5.2614 | 5.5059 | 0.9556 | 0.33928 |
| hispanic_rate | -1.8635 | 1.8807 | -0.9909 | 0.321758 |
| **Spatial Component** |  |  |  |  |
| Lambda | 0.28315 | 0.085763 | 3.3016 | 0.0049626 |
| Log likelihood | -1219.109 | N/A | N/A | N/A |
| Wald statistic | 10.9 | 0.085763 | 3.3016 | 0.00096146 |
| AIC | 2464.2 | N/A | N/A | N/A |

**Table 8:**
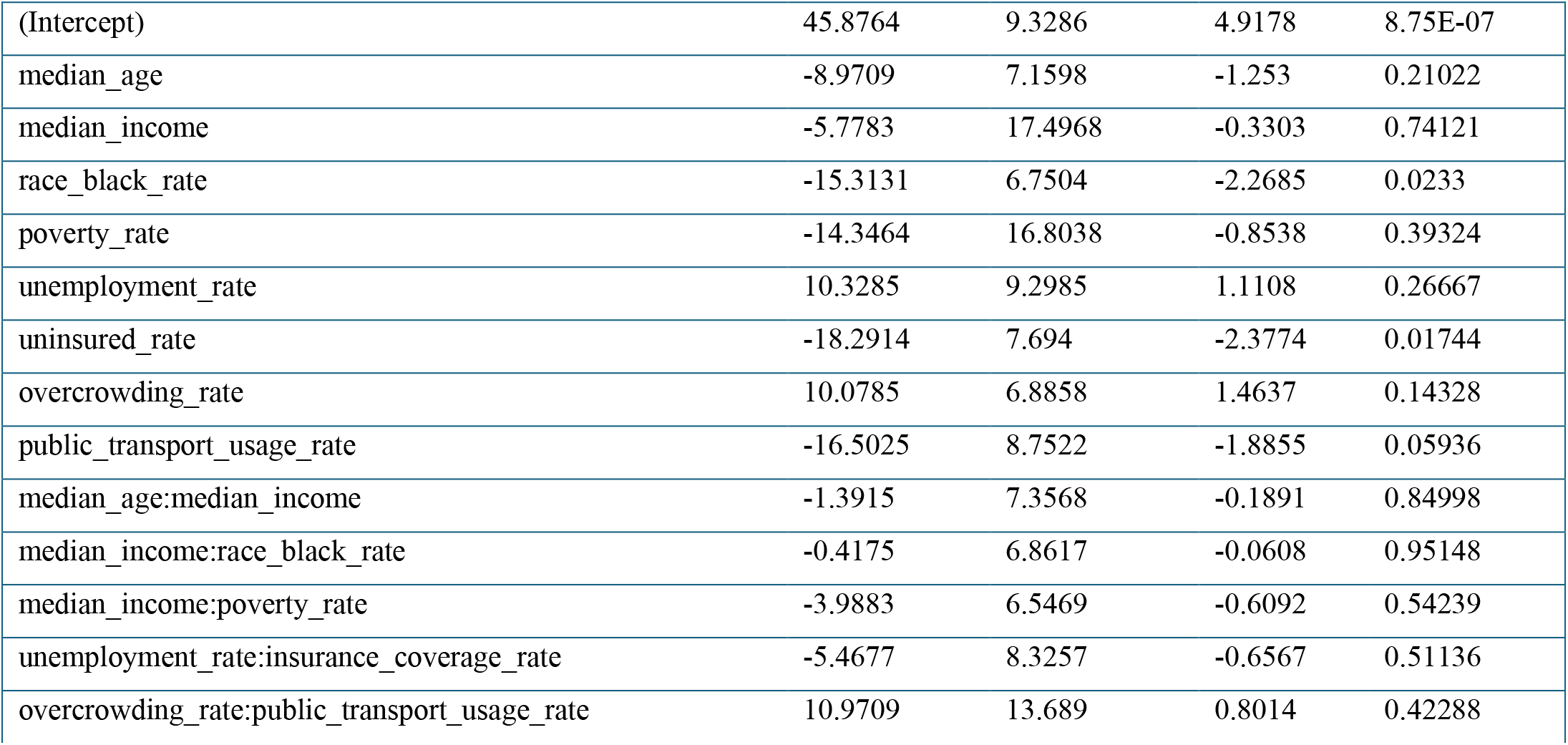
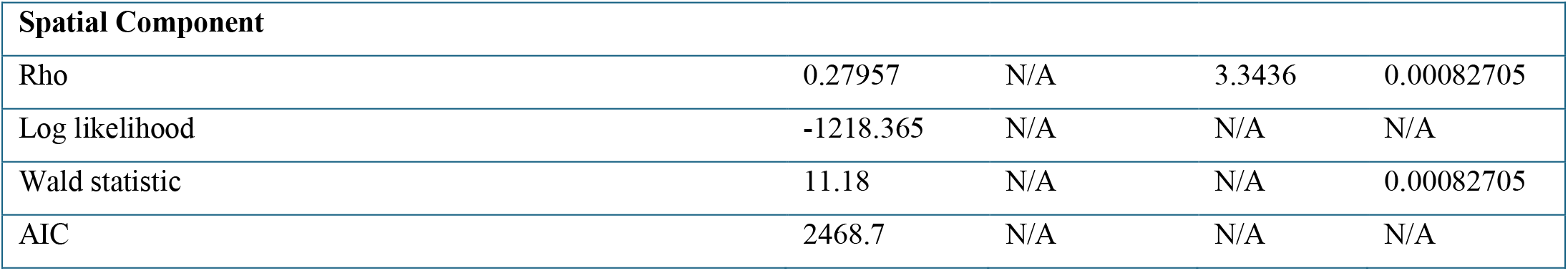
Spatial Lag with Interactions Model.

|  |  |  |  |  |
| --- | --- | --- | --- | --- |
| (Intercept) | 45.8764 | 9.3286 | 4.9178 | 8.75E-07 |
| median_age | -8.9709 | 7.1598 | -1.253 | 0.21022 |
| median_income | -5.7783 | 17.4968 | -0.3303 | 0.74121 |
| race_black_rate | -15.3131 | 6.7504 | -2.2685 | 0.0233 |
| poverty_rate | -14.3464 | 16.8038 | -0.8538 | 0.39324 |
| unemployment_rate | 10.3285 | 9.2985 | 1.1108 | 0.26667 |
| uninsured_rate | -18.2914 | 7.694 | -2.3774 | 0.01744 |
| overcrowding_rate | 10.0785 | 6.8858 | 1.4637 | 0.14328 |
| public_transport_usage_rate | -16.5025 | 8.7522 | -1.8855 | 0.05936 |
| median_age:median_income | -1.3915 | 7.3568 | -0.1891 | 0.84998 |
| median_income:race_black_rate | -0.4175 | 6.8617 | -0.0608 | 0.95148 |
| median_income:poverty_rate | -3.9883 | 6.5469 | -0.6092 | 0.54239 |
| unemployment_rate:insurance_coverage_rate | -5.4677 | 8.3257 | -0.6567 | 0.51136 |
| overcrowding_rate:public_transport_usage_rate | 10.9709 | 13.689 | 0.8014 | 0.42288 |

| Spatial Component |  |  |  |  |
| --- | --- | --- | --- | --- |
| Rho | 0.27957 | N/A | 3.3436 | 0.00082705 |
| Log likelihood | -1218.365 | N/A | N/A | N/A |
| Wald statistic | 11.18 | N/A | N/A | 0.00082705 |
| AIC | 2468.7 | N/A | N/A | N/A |

**Table 9:** Spatial Error with Interactions Model.

| Variable | Estimate | Std. Error | z value | Pr(>\|z\|) |
| --- | --- | --- | --- | --- |
| (Intercept) | 64.10389 | 9.32122 | 6.8772 | 6.10E-12 |
| `median_age` | -11.5548 | 7.43501 | -1.5541 | 0.12016 |
| `median_income` | -12.12399 | 18.02588 | -0.6726 | 0.50121 |
| `race_black_rate` | -19.00961 | 8.04292 | -2.3635 | 0.0181 |
| `poverty_rate` | -16.42961 | 16.85724 | -0.9746 | 0.32974 |
| `unemployment_rate` | 9.34689 | 9.85051 | 0.9489 | 0.34268 |
| `uninsured_rate` | -19.15362 | 7.64401 | -2.5057 | 0.01222 |
| `overcrowding_rate` | 7.23279 | 7.08871 | 1.0203 | 0.30757 |
| `public_transport_usage_rate` | -19.06628 | 9.8516 | -1.9353 | 0.05295 |
| `median_age:median_income` | -4.00621 | 7.38426 | -0.5425 | 0.58745 |
| `median_income:race_black_rate` | -0.34494 | 7.84032 | -0.044 | 0.96491 |
| `median_income:poverty_rate` | -4.62624 | 6.85432 | -0.6749 | 0.49971 |
| `unemployment_rate:insurance_coverage_rate` | -5.73692 | 8.83199 | -0.6496 | 0.51598 |
| `overcrowding_rate:public_transport_usage_rate` | 13.37656 | 12.34885 | 1.0832 | 0.27871 |
| Spatial Component |  |  |  |  |
| Lambda | 0.30463 | N/A | 3.6071 | 0.00030964 |
| Log likelihood | -1218.884 | N/A | N/A | N/A |
| Wald statistic | 13.011 | N/A | N/A | 0.00030964 |
| AIC | 2469.8 | N/A | N/A | N/A |

**Table 10:**
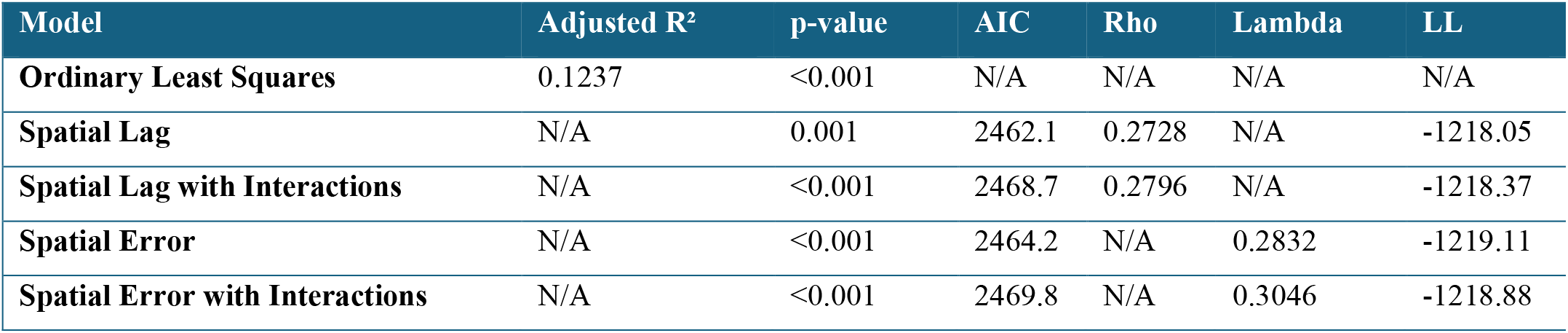
Overall Regression Model Results.

**Table 11:** Overall Regression Model Spatial Autocorrelation Tests.

| Model | Residual Autocorrelation Test |
| --- | --- |
| Ordinary Least Squares | Moran's I=0.125 (p=0.003) |
| Spatial Lag | LM=3.2509 (p=0.0714) |
| Spatial Lag with Interactions | LM=1.7322 (p=0.189) |
| Spatial Error | LR=7.893 (p=0.005) |
| Spatial Error with Interactions | LR=9.1198 (p=0.003) |

Similarly, the spatial error model with interactions was statistically significant (p < 0.001, table 8), and both race_black_rate and uninsured rate were significant predictors. Similar to the spatial lag model with interactions, none of the interaction terms were statistically significant predictors.

### Summary of Key Findings

1. **Uninsured Rate:** Higher rates of uninsured people were associated with a more negative trend in COVID-19 mortality rates. This suggests that counties with a larger uninsured population may have experienced a decline in mortality rates over time, potentially due to other factors like public health interventions or shifts in the pandemic’s impact across different population groups. However, it could also reflect delayed healthcare access or different timing of the pandemic’s severity in these areas.
2. **Black Population Rate**: A higher proportion of Black residents was negatively associated with COVID-19 mortality trends, indicating that counties with larger Black populations may have experienced declining mortality rates during the study period. This may reflect the disproportionate impacts at earlier stages of the pandemic, with mortality declining after heightened awareness and targeted interventions in these communities.
3. **Spatial Dependence (Rho)**: Both the spatial lag and spatial error models provided better fits to the data compared to the OLS model, highlighting the importance of accounting for spatial dependence when analyzing county-level mortality trends. These models indicated that spatial autocorrelation played a significant role in the mortality trends across counties, with nearby counties exhibiting similar trends.

## Discussion & Conclusion

This study investigated the relationships between socioeconomic status (SES) factors and COVID-19 mortality trends across U.S. counties, with a particular focus on spatially clustered counties identified using SatScan’s SVTT analysis. By applying spatial regression models, particularly the spatial lag model, we accounted for potential spatial autocorrelation, providing more robust insights into the regional dynamics of COVID-19 mortality trends.

The spatial lag model revealed three key findings:

### Uninsured Rate

Counties with higher rates of uninsured populations were significantly associated with more negative COVID-19 mortality trends, meaning mortality rates in these counties decreased over time. This counterintuitive result may reflect several factors. For instance, counties with low insurance coverage may have seen delayed impacts from the pandemic, or public health interventions, such as testing and vaccination, might have been intensified in areas with lower access to healthcare. Moreover, community-based support and delayed pandemic waves could also contribute to this observed decline in mortality.

### Black Population Rate

Another significant finding was the negative association between the proportion of Black residents and mortality trends. Counties with higher Black population rates experienced a decrease in mortality over time. This result suggests that targeted public health interventions aimed at reducing racial disparities—such as increased testing, vaccine distribution, and health outreach—were effective in mitigating the disproportionate impact of COVID-19 on Black communities, especially given the higher mortality rates early in the pandemic.

### Spatial Dependence

The spatial lag parameter (rho) was significant, underscoring the presence of spatial dependence in COVID-19 mortality trends. Counties with declining mortality rates tended to be geographically clustered, suggesting that public health measures, healthcare resources, and socioeconomic conditions in neighboring counties likely influenced each other. This finding emphasizes the importance of spatial relationships in shaping COVID-19 outcomes, highlighting the interconnectedness of regional responses to the pandemic.

#### Non-Significant Findings

Some SES variables, such as median age, median income, and poverty rate, did not emerge as significant predictors in the spatial lag model. This may be due to the overriding importance of healthcare-related factors, such as insurance coverage, during the pandemic. Although poverty and income are typically strong predictors of health outcomes, during a global health crisis like COVID-19, immediate access to healthcare services may have played a more direct role in reducing mortality than longer-term SES conditions.

#### Spatial Considerations & Model Performance

The spatial lag model outperformed the OLS model in terms of goodness of fit (as indicated by lower AIC values and significant spatial lag coefficients). By accounting for spatial dependence, the model was able to provide more accurate and reliable estimates of the associations between SES variables and COVID-19 mortality trends. Ignoring the spatial structure would have likely led to biased or inefficient estimates.

While residual autocorrelation was present in the OLS model, it was significantly reduced in the spatial lag model, highlighting the appropriateness of including spatial dependence in the analysis. This finding is consistent with the nature of the data, where counties sharing similar socioeconomic characteristics and public health responses influenced each other’s COVID-19 outcomes.

#### Limitations

A notable limitation of this study is the exclusion of counties not identified as clusters. While the decision to focus on clustered counties allows for the study of areas with significant COVID-19 mortality trends, it may limit the generalizability of the findings to counties not included in the clusters. Another limitation involves the reliance on county-level data, which may mask important within-county disparities, particularly in counties with large populations or diverse socioeconomic conditions.

## Data Availability

All data produced are available online at https://www.rwardrup.com/download/data-r-code-for-covid-19-spatial-clusters-ses/

## Compliance & Ethical Standards

### Funding

No funding was received for conducting this study.

### Ethics Approval

This study did not involve any human participants or animals, and thus ethics approval was not applicable.

### Consent to Participate

Not applicable.

### Consent for Publication

The authors provide consent for the publication of this study.

### Availability of Data and Material

The data used in this study are publicly available from CDC Wonder and ACS.

### Code Availability

https://www.github.com/minorsecond

## References

Desjardins, M. R., Hohl, A., & Delmelle, E. M. (2020). Rapid surveillance of COVID-19 in the United States using a prospective space-time scan statistic. International Journal of Health Geographics, 19(1), 1–8. 10.1016/j.apgeog.2020.102202

Gross, C. P., Essien, U. R., Pasha, S., Gross, J. R., Wang, S.-Y., & Nunez-Smith, M. (2021). Racial disparities in population-level COVID-19 mortality. Journal of General Internal Medicine, 36(2), 637–639. 10.1007/s11606-020-06081-w

Hawkins, R. B., Charles, E. J., & Mehaffey, J. H. (2020). Socio-economic status and COVID-19–related cases and fatalities. Public Health, 185, 129–134. 10.1016/j.puhe.2020.09.016

Kang, D., Choi, H., Kim, J.-H., & Choi, J. (2020). Spatial epidemic dynamics of the COVID-19 outbreak in China. International Journal of Infectious Diseases, 94, 96–102. 10.1016/j.ijid.2020.03.076

Kulldorff, M., Heffernan, R., Hartman, J., Assunção, R., & Mostashari, F. (2005). A Space–Time Permutation Scan Statistic for Disease Outbreak Detection. PLOS Medicine, 2(3), e59. 10.1371/journal.pmed.0020059

Millett, G. A., Jones, A. T., Benkeser, D., Baral, S., Mercer, L., Beyrer, C., Honermann, B., Lankiewicz, E., Mena, L., Crowley, J. S., Sherwood, J., & Sullivan, P. S. (2020). Assessing differential impacts of COVID-19 on Black communities. Annals of Epidemiology, 47, 37–44. 10.1016/j.annepidem.2020.05.003

Polyakova, M., Udalova, V., Kocks, G., Genadek, K., Finlay, K., & Finkelstein, A. (2021). Racial disparities in excess all-cause mortality during the COVID-19 pandemic. Proceedings of the National Academy of Sciences, 118(25), e2014746118. 10.1377/hlthaff.2020.02142

Shrestha, N., Shad MY., Ulvi, O., Khan, M. H., Karamehic-Muratovic, A., Nguyen, U.-S. D. T., … & Caceda, R. (2020). The impact of COVID-19 on globalization. One Health, 11, 100180. 10.1016/j.onehlt.2020.100180

Yancy, C. W. (2020). COVID-19 and African Americans. JAMA, 323(19), 1891–1892. 10.1001/jama.2020.6548

